# Characterizing physiological high-frequency oscillations using deep learning

**DOI:** 10.1101/2022.07.08.22277392

**Authors:** Yipeng Zhang, Hoyoung Chung, Jacquline P. Ngo, Tonmoy Monsoor, Shaun A. Hussain, Joyce H. Matsumoto, Patricia D. Walshaw, Aria Fallah, Myung Shin Sim, Eishi Asano, Raman Sankar, Richard J. Staba, Jerome Engel, William Speier, Vwani Roychowdhury, Hiroki Nariai

## Abstract

**Objective:** Intracranially-recorded interictal high-frequency oscillations (HFOs) have been proposed as a promising spatial biomarker of the epileptogenic zone. However, HFOs can also be recorded in the healthy brain regions, which complicates the interpretation of HFOs. The present study aimed to characterize salient features of physiological HFOs using deep learning (DL).

**Methods:** We studied children with neocortical epilepsy who underwent intracranial strip/grid evaluation. Time-series EEG data were transformed into DL training inputs. The eloquent cortex (EC) was defined by functional cortical mapping and used as a DL label. Morphological characteristics of HFOs obtained from EC (ecHFOs) were distilled and interpreted through a novel weakly supervised DL model.

**Results:** A total of 63,379 interictal intracranially-recorded HFOs from 18 children were analyzed. The ecHFOs had lower amplitude throughout the 80-500 Hz frequency band around the HFO onset and also had a lower signal amplitude in the low frequency band throughout a one-second time window than non-ecHFOs, resembling a bell-shaped template in the time-frequency map. A minority of ecHFOs were HFOs with spikes (22.9%). Such morphological characteristics were confirmed to influence DL model prediction via perturbation analyses. Using the resection ratio (removed HFOs/detected HFOs) of non-ecHFOs, the prediction of postoperative seizure outcomes improved compared to using uncorrected HFOs (area under the ROC curve of 0.82, increased from 0.76).

**Interpretation:** We characterized salient features of physiological HFOs using a DL algorithm. Our results suggested that this DL-based HFO classification, once trained, might help separate physiological from pathological HFOs, and efficiently guide surgical resection using HFOs.

## INTRODUCTION

Intracranially-recorded interictal high-frequency oscillations (HFOs) have been proposed as a promising spatial biomarker of the epileptogenic zone (EZ).^1, 2^ Animal and human studies have demonstrated the association between HFOs and brain tissue capable of generating seizures.^3-6^ Several retrospective studies have linked favorable post-surgical seizure outcomes to the resection of cortical sites showing interictal HFOs.^7-11^

However, despite the potential, HFOs can also be recorded in the healthy brain regions, which complicates the interpretation of HFOs when one attempts to guide resection using HFOs. Several recent studies, including a large multicenter prospective study, failed to correlate the removal of HFO-generating brain regions with postoperative seizure freedom; some patients became seizure-free despite part of the brain regions generating HFOs being preserved.^12, 13^ An ongoing clinical trial utilizing HFOs in electrocorticography to guide resection has excluded enrollment of occipital lobe epilepsy, due to abundant physiological HFOs in the visual areas.^14^ The current impasse is that there are no methods to separate pathological from physiological HFOs.

Within the field of artificial intelligence, machine learning can bridge statistics and computer science to develop algorithms to complete tasks by exposure to meaningful clinical data without explicit instruction. Indeed, machine learning has been successfully applied to the problem of classifying HFOs based on a priori manual engineering of event-wise features, which includes: linear discriminant analysis,^15^ support vector machines,^16, 17^ decision trees,^18^ and clustering.^19^ More recently, the deep learning (DL) framework has been adopted, which directly works with raw data (avoiding any a priori feature engineering) and yields better performance in the field of neuroimaging.^20^ Leveraging DL’s revolutionary success in the field of computer vision using Convolutional Neural Networks (CNNs), prior studies explored the use of CNNs in EEG analysis, especially converting one-dimensional EEG signal into a two-dimensional image for CNNs input.^21-23^ The previous DL approaches conducted the HFO classification in a supervised manner, requiring human annotated labels which constrains the spectrum of usage of their methods, especially the needs of human expert labeling. In the context of medical image analysis, recent work has shown that optimized model architectures and loss functions could mitigate data labeling errors, thus making the DL framework even more applicable.^24^ Our recent work demonstrated that using the channel resection status as DL labels, a novel weakly-supervised DL algorithm characterized HFOs generated by the epileptogenic zone.^25^

The present study aimed to characterize physiological HFOs, using functional cortical mapping results as DL training labels. We leveraged results from functional cortical mapping by stimulation and gamma-related language mapping in children with neocortical medication-resistant epilepsy who underwent intracranial EEG monitoring with a grid/strip approach. We characterized salient features of physiological HFOs represented by the eloquent cortices (ecHFOs) through the DL algorithm and also investigated if removal of cortical regions with HFOs excluding ecHFOs (non-ecHFOs, deemed pathological HFOs) correlated with postoperative seizure-freedom.

## METHODS

### Patient cohort

This was a retrospective cohort study. Children (below age 21) with medically refractory epilepsy (typically with monthly or greater seizure frequency and failure of more than three first-line anti-seizure medications) who had intracranial electrodes implanted for the planning of epilepsy surgery with anticipated cortical resection with the Pediatric Epilepsy Program at UCLA were consecutively recruited between August 2016 and August 2018. Diagnostic stereo-EEG evaluation (not intended for resective surgery) was excluded.

### Standard protocol approvals, registrations, and patient consents

The institutional review board at UCLA approved the use of human subjects and waived the need for written informed consent. All testing was deemed clinically relevant for patient care, and also all the retrospective EEG data used for this study were de-identified before data extraction and analysis. This study was not a clinical trial, and it was not registered in any public registry.

### Patient evaluation

All children with medically refractory epilepsy referred during the study period underwent a standardized presurgical evaluation, which—at a minimum—consisted of inpatient video-EEG monitoring, high resolution (3.0 T) brain magnetic resonance imaging (MRI), and 18 fluoro-deoxyglucose positron emission tomography (FDG-PET), with MRI-PET co-registration.^26^ The margins and extent of resections were determined mainly based on seizure onset zone (SOZ), clinically defined as regions initially exhibiting sustained rhythmic waveforms at the onset of habitual seizures. In some cases, the seizure onset zones were incompletely resected to prevent an unacceptable neurological deficit.

### Subdural electrode placement

Macroelectrodes, including platinum grid electrodes (10 mm intercontact distance) and depth electrodes (platinum, 5 mm intercontact distance), were surgically implanted. The total number of electrode contacts in each subject ranged from 40 to 128 (median 96 contacts). The placement of intracranial electrodes was mainly guided by the results of scalp video-EEG recording and neuroimaging studies. All electrode plates were stitched to adjacent plates, the edge of the dura mater, or both, to minimize the movement of subdural electrodes after placement.

### Acquisition of three-dimensional (3D) brain surface images

We obtained preoperative high-resolution 3D magnetization-prepared rapid acquisition with gradient echo (MPRAGE) T1-weighted image of the entire head. A FreeSurfer-based 3D surface image was created with the location of electrodes directly defined on the brain surface, using post-implant computed tomography (CT) images.^27^ In addition, intraoperative pictures were taken with a digital camera before dural closure to enhance the spatial accuracy of electrode localization on the 3D brain surface. Upon re-exposure for resective surgery, we visually confirmed that the electrodes had not migrated compared to the digital photo obtained during the electrode implantation surgery.

### Intracranial EEG (iEEG) recording

Intracranial EEG (iEEG) recording was obtained using Nihon Kohden Systems (Neurofax 1100A, Irvine, California, USA). The study recording was acquired with a digital sampling frequency of 2,000 Hz, which defaults to a proprietary Nihon Kohden setting of a low frequency filter of 0.016 Hz and a high frequency filter of 600 Hz at the time of acquisition. For each subject, separate 10-minute and 90-minute EEG segments were selected at least two hours before or after seizures, before anti-seizure medication tapering, and before cortical stimulation mapping, which typically occurred two days after the implant. All the study iEEG data were part of the clinical EEG recording.

### Functional cortical mapping

Cortical stimulation was performed as part of clinical management to define regions of the eloquent cortices (EC) to help guide resections. A pulse train of repetitive electrical stimuli was delivered to neighboring electrode pairs, with a stimulus frequency of 50 Hz, pulse duration of 300 µs, and train duration ranging up to 5 seconds (sensorimotor or visual mapping) or 7 seconds (language mapping). Stimulus intensity ranged from 1 mA to 13 mA. Seven out of 19 patients also underwent auditory and picture-naming tasks to supplement cortical stimulation mapping. In short, patients were instructed to overtly verbalize an answer to a given auditory question or name a picture. Each EEG trial data was transformed into the time-frequency domain using complex demodulation via BESA software (BESA GmbH, Germany). The iEEG signal at each channel was assigned an amplitude (a measure proportional to the square root of power) as a function of time and frequency (in steps of 10 ms and 5 Hz). The time-frequency transform was obtained by multiplication of the time-domain signal with a complex exponential, followed by a band-pass filter. iEEG traces were aligned to: (i) stimulus (question) onset; (ii) stimulus offset; and (iii) response (answer) onset. We determined whether the degree of such gamma-augmentation reached significance using studentized bootstrap statistics followed by Simes’ correction.^28, 29^ Sites surviving correction showing significant gamma-augmentation spanning (i) at least 20-Hz in width and (ii) at least 20-ms in duration were defined as ‘language-related gamma sites’.^30^ Further methodological details of the gamma mapping are described in the prior studies.^31^

### Automated detection and classification of HFOs

A customized average reference was used for the HFO analysis, with the removal of electrodes containing significant artifacts.^26, 32^ Candidate interictal high frequency oscillations (HFOs) were identified by an automated short-term energy detector (STE).^33, 34^ This detector considers HFOs as oscillatory events with at least six peaks and a center frequency occurring between 80-500 Hz. The root mean square (RMS) threshold was set at five standard deviations (SD), and the peak threshold was set at three SD. The HFO events are segments of EEG signals with durations ranging from 60 to 200 ms.^34^ The candidate HFOs were evaluated by our previously validated algorithm (recall = 98.0%, precision = 96.1%, F1 score = 96.8% against human experts validation) to reject artifacts for further analyses.^25^ For simplicity, we use HFO in the later paragraphs to represent the HFO after the artifact rejection.

### Weakly supervised deep learning using functional cortical mapping results as labels

The general workflow of the DL training and inference was shown in the flowchart (**Figure 1A**). The channel-wise annotations include behavior (positive behavioral changes with cortical stimulation mapping or language-related gamma sites), SZ/AD (channels with seizures or afterdischarges with cortical stimulation mapping), spike (cortical sites showing spontaneous interictal spikes), and none (no behavioral changes with mapping, SZ/AD, and spikes). We hypothesize that a morphologically distinct class of HFOs generated by EC, representative as physiological HFOs (defined as ecHFOs). We expect a large percentage of the HFOs in the behavioral channels are ecHFOs. However, such channels could also have HFOs that are morphologically distinct from ecHFOs, since each channel also picks up neuronal activities generated by other physiological or pathological processes. We refer to such complementary HFOs as non-ecHFOs. Since the physiological processes generating ecHFOs are also present in different regions of the brain, for example, the unstimulated brain region, we expect non-behavioral channels to also have ecHFOs. However, as there is no ground truth annotation for an ecHFO event, we relied on channel-level annotations acquired from clinical experiments. Specifically, positive labels are assigned to HFOs from channels that only have behavior responses, excluding spike and SZ/AD channels, and negative labels are assigned to all other HFOs. When an HFO is from a channel with both behavior response and pathological (SZ/AD or Spike) response, we excluded them from the training set, as the distribution of ecHFOs and non-ecHFOs might be comparable. Clearly, such labeling has errors. However, if ecHFOs have distinct morphological signatures, then we could utilize the generalization power of the neural network to distill the ecHFO from this weak supervision.

**Figure 1:**
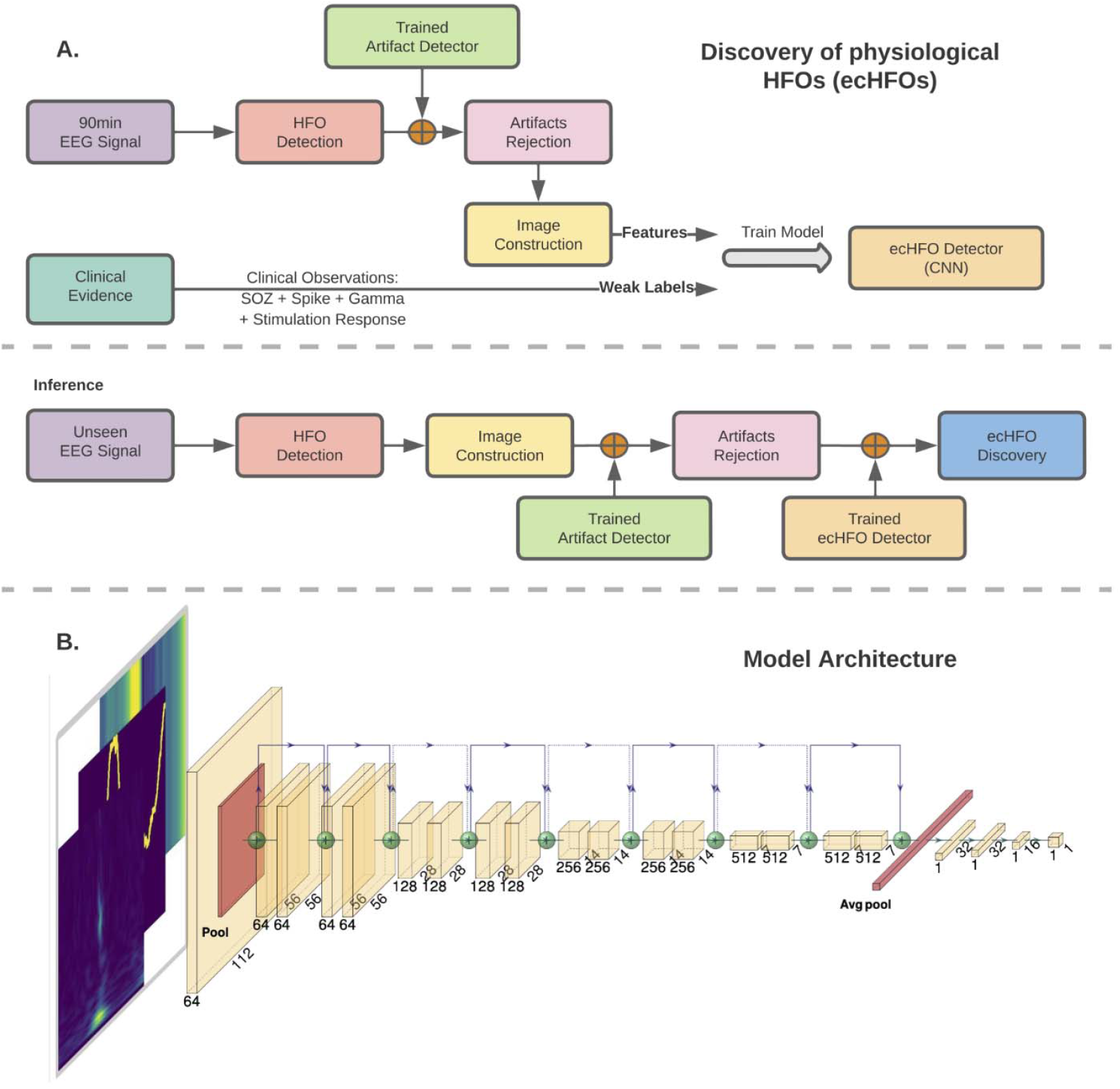
Processing workflow. **(A)**: The detected HFO (by STE detector) is first filtered by the artifact detector. The constructed image features along with the channel-wise clinical information are fed into the convolutional neural network to train the model. **HFO feature representation and model architecture (B)**: We captured t e time-frequency domain features as well as signal morphology information of the HFO window via three images. The time-frequency plot (scalogram) was generated by continuous Gabor Wavelets ranging from 10Hz to 500Hz. The EEG tracing plot was generated on a 2000 × 2000 image by scaling the time-series signal into the 0 to 2000 range to represent the EEG waveform’s morphology. The amplitude-coding plot was generated to represent the relative amplitude of the time-series signal: for every time point, the pixel intensity of a column of the image represented the signal’s raw value at that time. These three images were resized into the standard size (224 × 224), serving as the input to the neural network. We used ResNet 18 with a modified output layer for binary classification. The weights in the convolution layers are frozen and serve as feature extractors in the convolutional neural network.

### Model architecture and Feature representation of HFOs

We adopt the same CNN architecture as the prior study because it has demonstrated excellent performance in classifying HFOs.^25^ A one-second window represented each HFO, centered on the HFO (0 ms). Then three images are constructed to capture the time-frequency domain features as well as signal morphology information of the HFO window. Then these three images were resized into the standard size (224 × 224), serving as the input to the neural network (**Figure 1B**).

### Model training

We used 90 minutes of data from each patient for training. Since our task is binary classification, we use binary cross-entropy as the loss function, L=−[y · log(x)+(1−y)·log(1−x)], where y is the label (1 for ecHFO, 0 for non-ecHFO [ecHFOs were subtracted from total HFOs]), and x is the model output ecHFO probability. In training, we adopt stratified sampling to balance the data distribution in different labels. The Adam optimizer was adopted with a learning rate of 0.0001. The training was conducted using 25 epochs (training iterations), and validation loss was plotted with respect to the number of epochs completed. For selecting the best model across epochs, we picked the model corresponding to the balanced global minima over 25 iterations, i.e., the highest balanced accuracy (the averaged recall for each class). In order to fully explore the performance of the model, five-fold cross-validation was conducted using the pooled data across the full patient cohort. For each fold, 20% of the dataset was selected as the test set, 70% was selected as the training set, and the remaining 10% was used for validation.

### Performance analysis framework

Since the model was evaluated by using five-fold cross-validation, there were five distinctive models trained by different folds of data. For the downstream analysis, if an HFO is from a channel with both behavior response and SZ/AD response, the classification of these HFOs is determined by the mean of the probabilities from five models. Otherwise, the classification of the HFO is determined by the model where this specific HFO is in the test set.

### Characterization of ecHFOs

We adopted the same procedure as the prior study to determine whether the time-frequency scalogram of ecHFOs differed from that of non-ecHFOs, that is, conducting a t-test across all pixels in the time-frequency scalogram (**Figure 2A**). The null hypothesis is that the values within the scalogram of ecHFO are not lower than those of non-ecHFO. We summarized the results via a histogram of the change in probabilities for each patient across all predicted non-ecHFOs. Additionally, a one-tailed t-test comparing values of ecHFOs and non-ecHFOs was performed on the change of output probability score to ensure that the change was significant and generalized well at the population level (**Figure 2B**).

**Figure 2:**
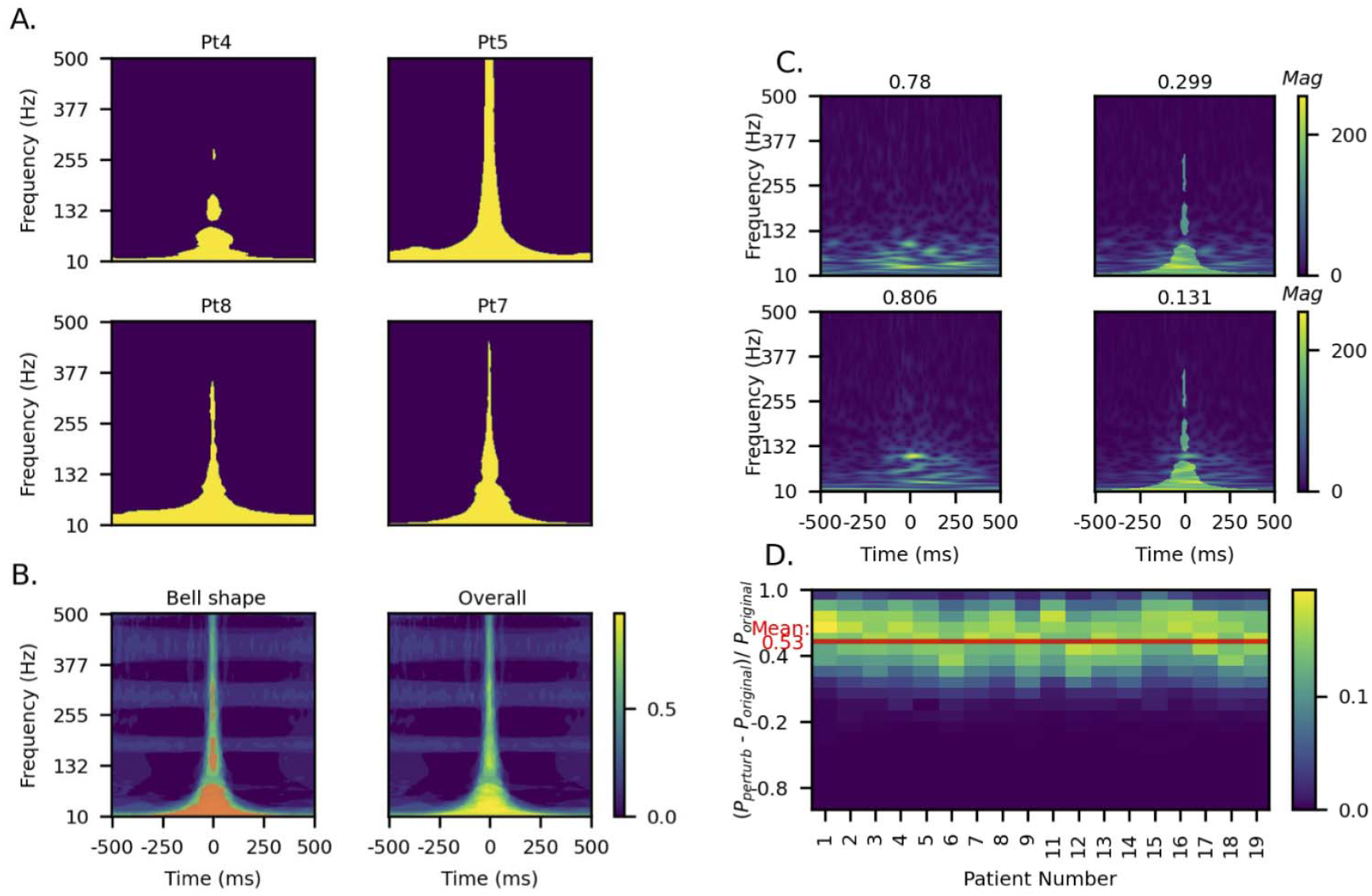
Characteristics in the time-frequency plot of ecHFOs against non-ecHFOs. (A) The time-frequency plot characteristics of the eloquent cortex and non-eloquent cortex HFOs for Pt 4, 5, 7, and 8. The yellow-colored regions in the figure stood for the pixels, where the power spectrum of ecHFOs is statistically lower than (P-value below 0.05 from the one-tailed t-test) non-ecHFOs. The figure showed one set of clearly interpretable distinguishing features between ecHFOs and non-ecHFOs: the ecHFOs generally have lower power at lower frequencies during the HFO event (center part along the time axis), Panel (B-top) was generated by taking the average of the individual binary images from each of the 18 patients. It showed the distinguishing features are also significant at the population level. The feature can be assembled as a “Bell-Shape” if we take the region > 0.7 in the plot averaging all of the time-frequency plot characteristics (red color). (B-bottom). (C) The model’s response to the Bell-shaped perturbation on the time-frequency plot. We provide two examples of perturbatio for ecHFO events in Pt 3. Each row presents one example and the first column indicates the original time-frequency plot while the second indicates the perturbed time-frequency plot based on the Bell Shape perturbation. The prediction value of the model changed from 0.753 (therefore originally predicting it as non-ecHFO) to 0.1 (thus a change of 0.653), implying that the perturbed HFO would correspond to a non-ecHFO. (D) The change in model confidence in population level. Each column (along the y-axis) is a histogram of the percentage of change in confidence for one distinct patient. It shows the frequency distribution of confidence changes after adding the bell shape perturbation to the time-frequency plot to all classified ecHFOs for the given patient. The change in confidence level is significant, with an average of 53% with a 95% confidence interval [52.8, 53.2] noted as the solid red line in the histogram. HFO = high-frequency oscillation; ecHFOs = eloquent cortex HFOs; non-ec = non-eloquent cortex HFOs.

### Interpretability analysis of ecHFOs

Perturbation on time-frequency plot: We adopted a similar perturbation analysis as prior work based on the characterization of ecHFO in the time-frequency plot (**Figure 2C**). We summarized the results via the histogram of the percentage of change in probabilities for each patient across all the predicted ecHFOs (**Figure 2D**). A one-tailed t-test was also performed on the change of that output probability score to ensure that the change was significant and generalized well at the population level.

Perturbation on amplitude coding plot: We adopted a data-driven approach to discover the effect of introducing a spike in the amplitude coding plot (**Figure 3**). As in our prior work, we built a detector to capture HFO with a spike (spk-HFO) trained on expert annotations (86.5% accuracy with an F1 score of 80.8%).^25^ The spk-HFO detector was applied to our data and predicted whether each HFO event was an spk-HFO or not. For all detected spk-HFOs in each patient, we separated these HFOs into upward spikes and downward spikes. The upward (downward) spikes are defined as spk-HFO events, with the average amplitude in the center 20-ms duration being greater (smaller) than the average amplitude in the peripheral 250 ms. We then created upgoing and downgoing spike templates by taking the average of all tracings. The corresponding amplitude coding plots of the upgoing and downgoing spike templates are built for perturbing the amplitude coding plots for each patient (**Figure 3A, C**). This perturbation is performed by adding two times the template to the amplitude coding plot of each predicted ecHFO and measuring the change in model confidence. We summarized the results via the histogram of the percent change in probability for each patient across all the predicted ecHFOs (**Figure 3B, D**). A one-tailed t-test was performed on the change of output probability to evaluate whether the change was significant and generalized well at the population level.

**Figure 3:**
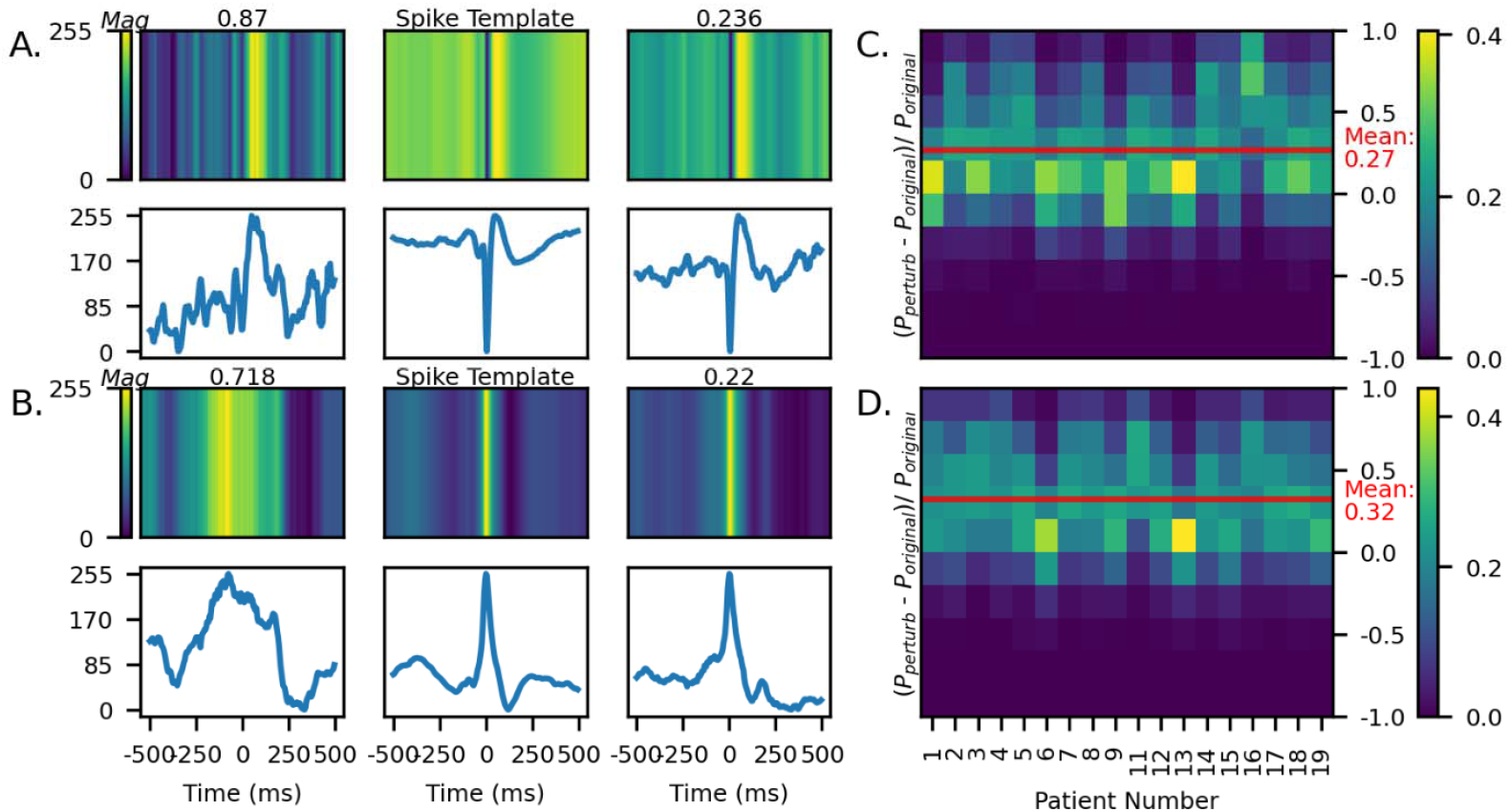
The model’s responses to injecting a spike-like feature into the amplitude-coding plot. Examples of introducing a downgoing (A) and upgoing (C) spike feature into classified non-ecHFO events. These demonstrate that on the introduction of a spike-like perturbation, the model predicts higher confidence towards non-ecHFOs (B, D). Subfigure A shows the amplitude encoding image before perturb, spike template, and the after-perturb (top row), and the corresponding time-series signal with downgoing spike perturbation (bottom row). Similarly, subfigure C shows the same information on a different classified ecHFO but with upgoing spike perturbation. For each patient, we compute a histogram for the distribution of the change in confidence (B). The same steps are repeated for upgoing spike perturbation, and the results are shown in (D). The percentage of change in confidence for both up-and-downgoing spike perturbation is significantly greater than zeros, with means downgoing: 0.268 (95% confidence interval [0.2633, 0.2723]) and upgoing: 0.320 (95% confidence interval [0.3157, 0.3237]), which are noted as solid red lines in each histogram. HFO = high-frequency oscillation; ecHFOs = eloquent cortex HFOs; non-ecHFO = non-eloquent cortex HFOs.

### Time Domain Characteristic ecHFO

We reported three time-domain characteristics of the predicted ecHFOs and non-ecHFOs, which are amplitude, duration, and max frequency. The duration is defined as the time duration of the HFO predicted by the RMS HFO detector **(Figure 4**). The amplitude is defined as the maximum absolute value within the HFO interval, and the max frequency is defined as the largest frequency component within a window (0.2 seconds before and after) around the center of the HFO. We plotted the normalized histogram to compare the distribution of these three characteristics.

**Figure 4:**
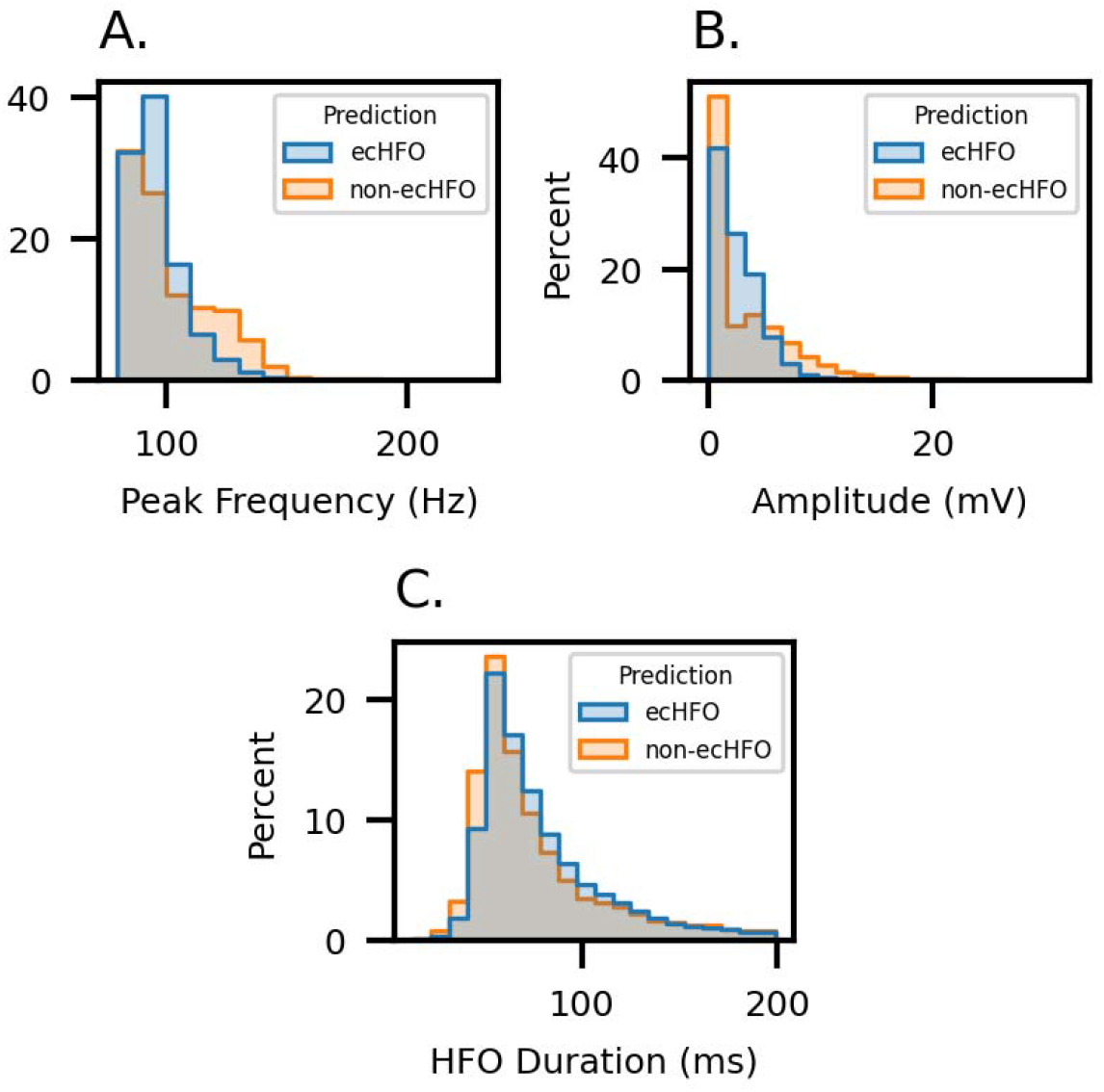
Traditional feature characterization of ecHFO and Non-ecHFO: The normalized histogram of peak frequency (A), amplitude (B), and duration (C) of predicted ecHFO. (A) The ecHFO generally has a lower peak frequency than the non-ecHFO (p-value < 0.001). (B) The ecHFO generally has a smaller amplitude than the non-ecHFO (p-value < 0.001). (C) The ecHFO generally has the trend of having a longer HFO than the non-ecHFO (p-value = 0.07). However, there are no clear decision boundaries that can be drawn to clearly discriminate between ecHFO and non-ecHFO using each of these traditional features.

### Comparison of resection ratios of HFOs to postoperative seizure outcomes

We estimated the probability of each patient’s surgical success (for the 14 patients who underwent resective surgery with known seizure outcomes at 24 months) based on the resection ratio of HFOs (number of resected HFOs/number of detected HFOs) as a classifier. We constructed the receiver operating characteristic (ROC) curve and calculated the area under the curve (AUC) values in the resection ratio of unclassified HFOs and non-ecHFOs. Determination of the channel resection status (resected vs. preserved) was determined based on intraoperative pictures (pre-and post-resection) and also on post-resection brain MRI, based on discussion among a clinical neurophysiologist (HN), neurosurgeon (AF), and radiologist (SN). A multiple logistic regression model incorporating the resection ratio of non-ecHFOs and complete resection of SOZ was also created. The surgical outcomes were determined 24 months after resection as either seizure-free or not seizure-free.

### Statistical analysis

Above mentioned statistical calculations were carried out using Python (version 3.7.3; Python Software Foundation, USA). The deep neural network was developed using PyTorch (version 1.6.0; Facebook’s AI Research lab). Quantitative measures are described by medians with interquartile or means with standard deviations. Comparisons between groups were performed using chi-square for comparing two distributions and Student’s t-test for quantitative measures (in means with standard deviations). All comparisons were two-sided and significant results were considered at p < 0.05 unless stated otherwise. Specific statistical tests performed for each experiment were described in each section.

### Data sharing and availability of the methods

Anonymized EEG data used in this study are available upon reasonable request to the corresponding author. The python-based code used in this study is freely available at (https://github.com/roychowdhuryresearch/HFO-Classification). One can train and test the deep learning algorithm from their data and confirm our methods’ validity and utility.

## RESULTS

### Clinical information (patient characteristics)

There were 19 patients (10 females) enrolled during the study period. The median age at surgery was 14 years (range: 3-20 years). The median electrocorticography monitoring duration was 4 days (range: 2-14 days), and the median number of seizures captured during the monitoring was 8 (IQ range: 4-25). There were 15 patients who underwent resection, and 14 patients provided postoperative seizure outcomes at 24 months (9 of 14 became seizure-free). Due to unknown postoperative seizure outcomes, patient #10 was removed from the further HFO analyses. Details of patients’ clinical information are listed in Table 1.

**Table 1.**
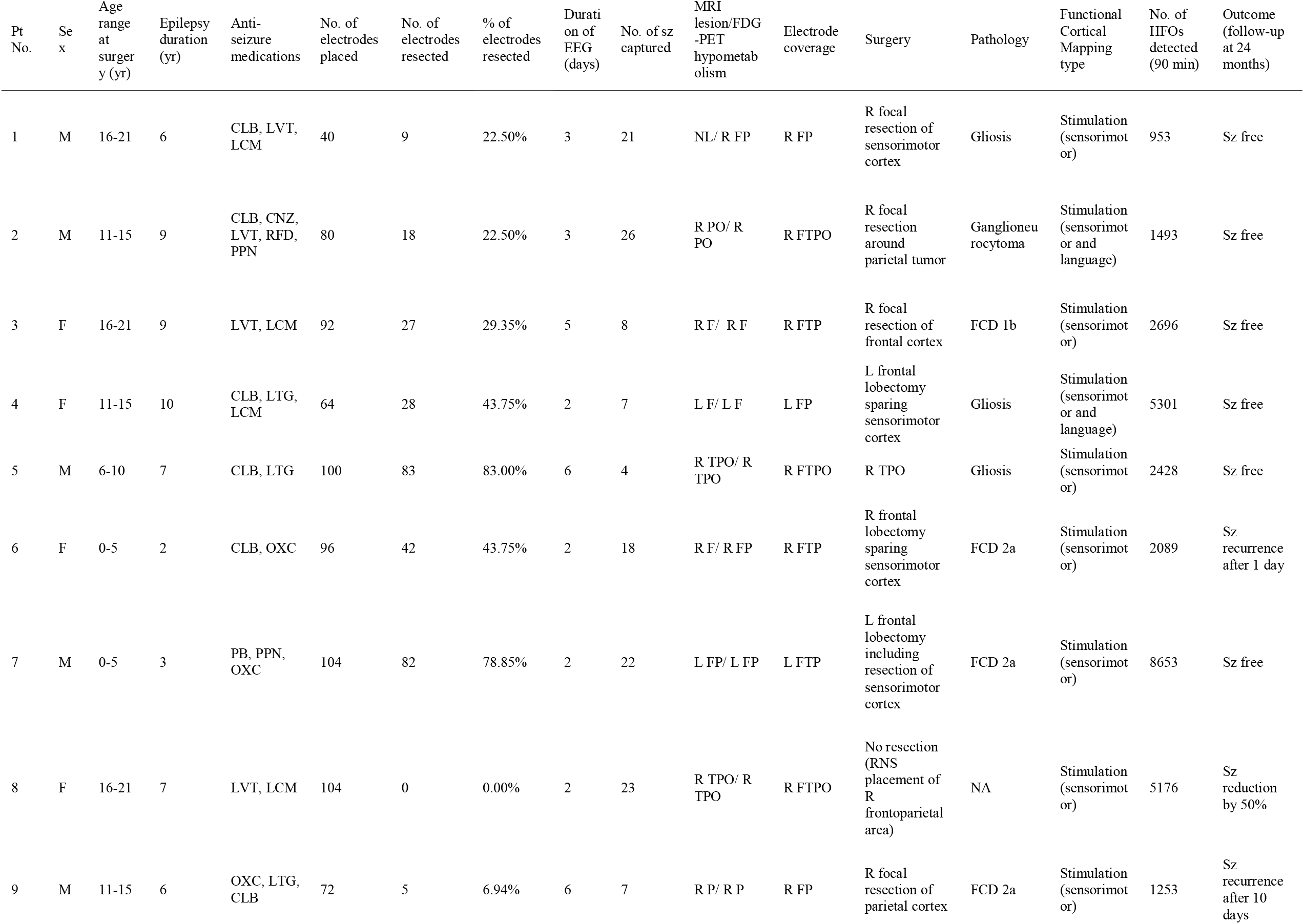
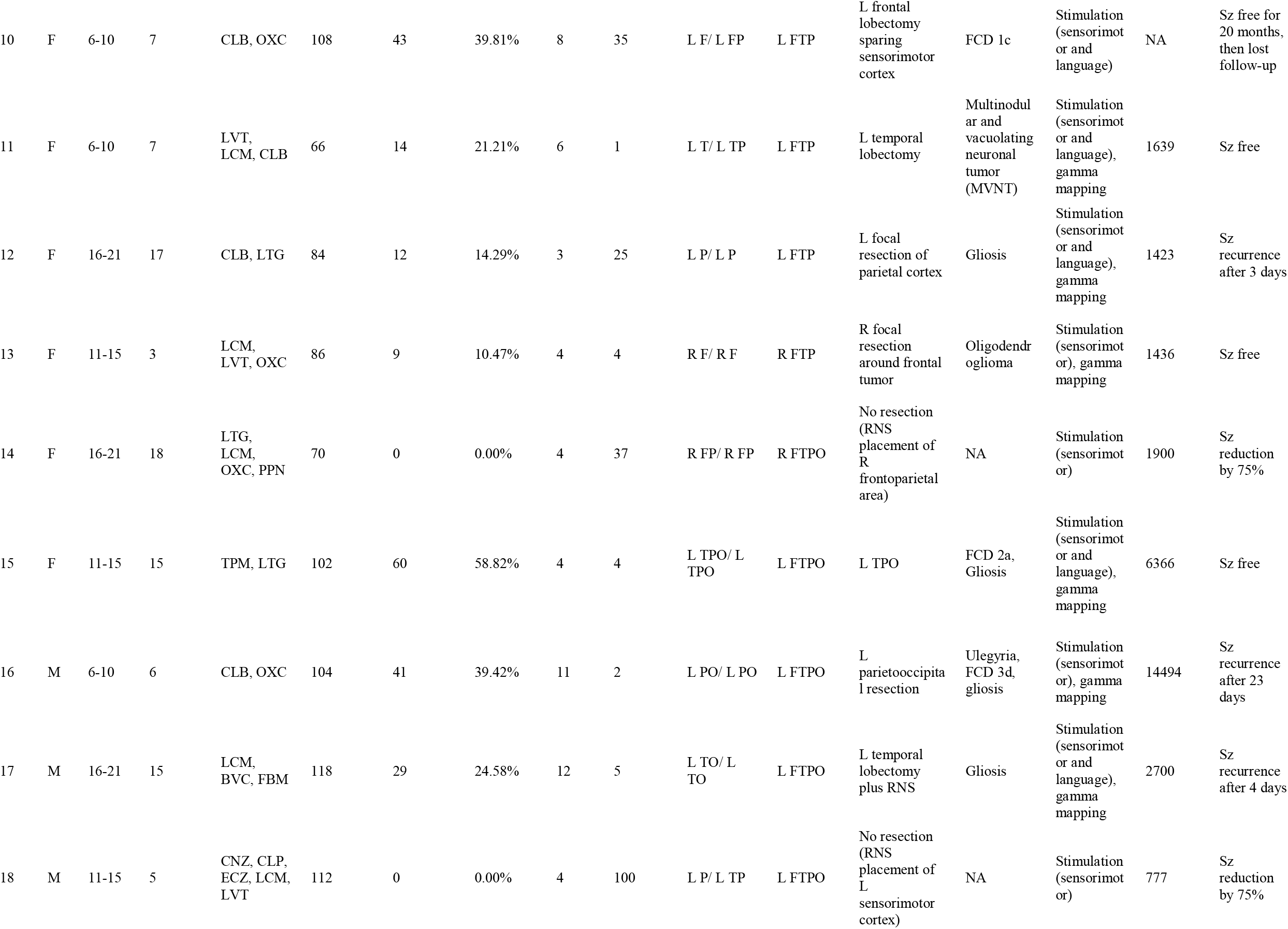
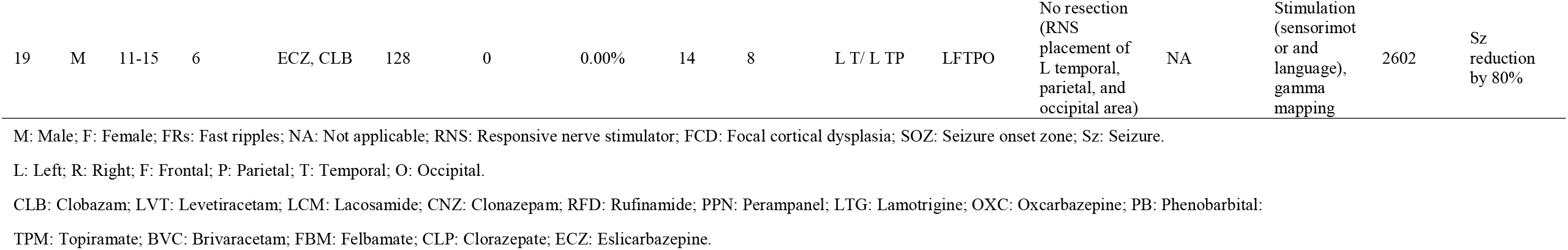
Cohort characteristics.

### Interictal HFO detection

A total of 63379 HFO events were detected (median 2258.5 events per patient) in 90-minute EEG data from the 18 patients (patient #10 was removed for subsequent HFO analysis due to lack of seizure outcomes). There were 15111 HFOs recorded from the stimulated channels (48268 HFOs from non-stimulated channels). There were 7173 HFOs obtained from behavior channels (positive behavioral responses from cortical stimulation mapping and/or gamma mapping), and “behavior only” channels (behavior channels without spontaneous spikes or SZ/AD) exhibited 3822 HFOs. There were 5317 HFOs obtained from “none” channels (no spontaneous spikes and no behavioral responses with mapping). After the DL training, we noted that 22.9% (4547/19816) of the ecHFOs were HFOs with spikes, and 77.1% (30647/43563) of non-ecHFOs were HFOs with spikes (p < 0.0001, a chi-square test).

### Time-frequency plot characteristics of ecHFO and non-ecHFOs

The analysis of the time-frequency map demonstrated that ecHFOs had lower amplitude throughout the frequency band, including both ripples (80-250 Hz) and fast ripples (250-500 Hz) around the center point (0 ms, where HFOs were detected) than non-ecHFOs (**Figure 2A**). We inspected the t-test map and selected regions greater than 0.7 to define the salient characteristic difference between ecHFOs and non-ecHFOs. The selected region resembled a bell-shaped (**Figure 2B**).

### Perturbation analyses

By utilizing the bell-shaped template found in the time-frequency map, we observed that the bell-shaped perturbation on the time-frequency plot significantly increased the model prediction probability towards non-ecHFOs (mean probability increase was 0.532 [95% confidence interval: 0.528–0.532], p < 0.001) (**Figure 2C, D**). Furthermore, we analyzed the effect of introducing a spike-like shape in the amplitude-coding plot. By introducing a downgoing or an upgoing spike in an ecHFOs event, the model confidence increased towards a non-ecHFO event (**Figure 3A, C**). On the population level, the prevalent probability increase in ecHFO events among all patients (**Figure 3B, D**) demonstrated the non-trivial model response by introducing a spike in the time domain (mean probability increase of 0.268 for a downgoing spike introduction [95% confidence interval: 0.2633–0.2723], and 0.320 for an upgoing spike introduction [95% confidence interval: 0.3157–0.3237], both with p< 0.001).

### Feature characterization of ecHFOs vs. non-ecHFOs

We plotted the histogram of the peak frequency, amplitude, and duration of both ecHFO and non-ecHFO, respectively (**Figure 4**). The ecHFOs exhibited a smaller amplitude (p-value < 0.001), a lower max frequency (p-value < 0.001) and a trend of shorter HFO length (p-value = 0.07) than non-ecHFO. However, there was no decision boundary to clearly discriminate between ecHFO and non-ecHFO using each of these traditional features.

### Inference of channel characteristics

After the model was trained, it assigned one prediction label for all of the HFOs in each channel, and we plotted the ecHFO ratio (proportion of ecHFOs divided by the number of all HFOs) and the model confidence distribution towards ecHFOs in different channels (**Figure 5**). In channels with a physiological response (only behavior responses with functional mapping), the ecHFO ratio was high, and confidence scores were skewed towards ecHFOs. Channels with a pathological response (spike and SZ/AD with stimulation) showed a lower ecHFO ratio, and confidence scores were skewed towards non-ecHFOs. Channels with no behavioral responses showed the confidence scores skewed towards non-ecHFOs. Notably, channels with both properties, in which both behavioral responses and spikes or/and SZ/AD, model confidence showed a uniform distribution, suggesting the presence of both ecHFOs and non-ecHFOs.

**Figure 5:**
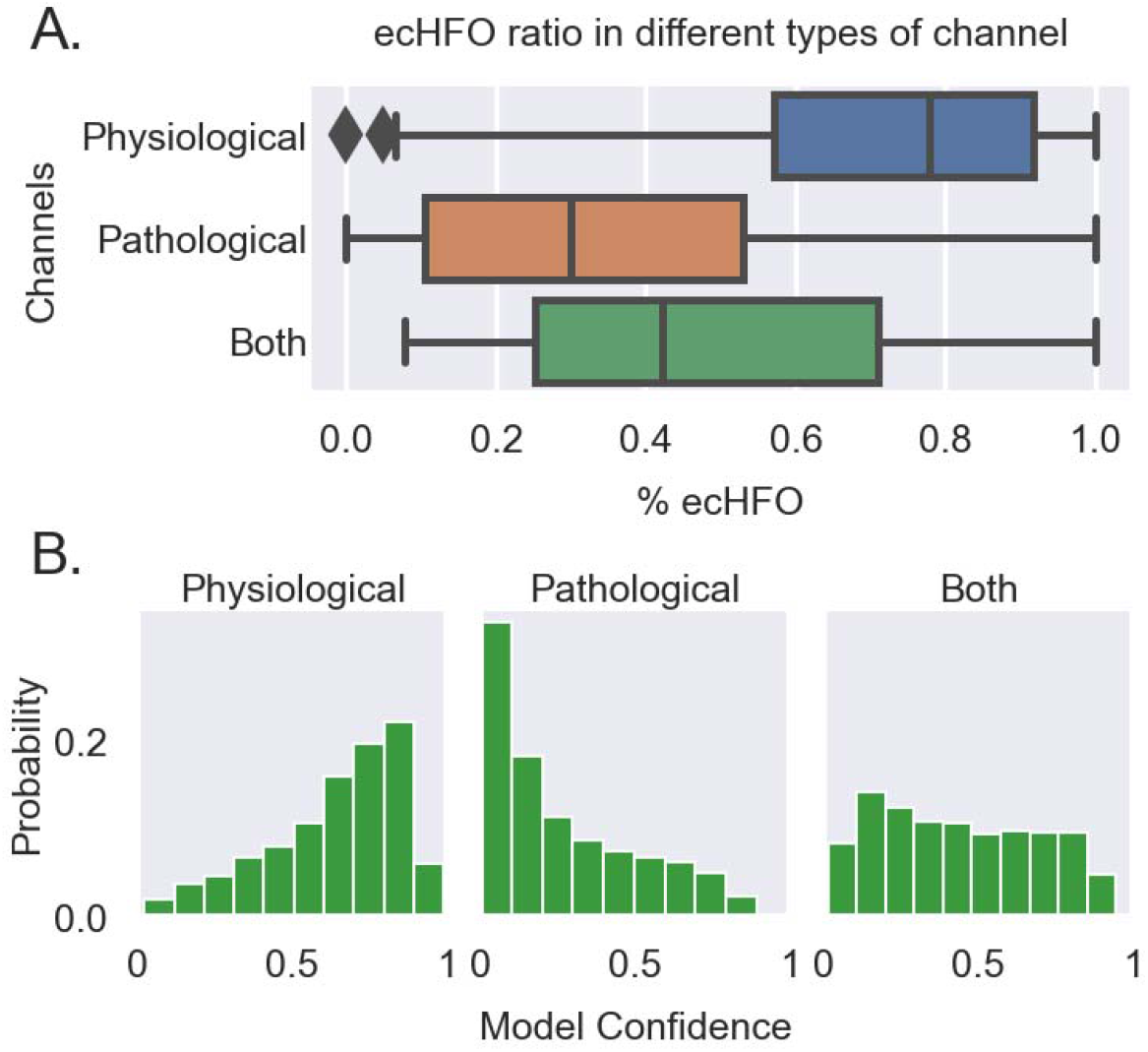
Inference of channel characteristics: (A) The ratio of ecHFOs (eHFOs/total HFOs) in different types (physiological: behavior or gamma only; pathological: SZ/AD or spike only; both: both physiological and pathological) of channels from all patients (n=14) is plotted in box plots. The percentage of ecHFOs was higher in physiological channels than that in pathological channels. (B) The model confidence distribution of each individual eHFOs in channels with Physiological, Pathological, and both categories are shown. The distribution in both channels is closer to a uniform distribution.

### Clinical inference using postoperative seizure outcomes

We created the ROC curves using the HFO resection ratio to predict postoperative seizure freedom at 24 months (n = 14) (**Figure 6**). Using the resection ratio of HFOs showed acceptable prediction performance (AUC= 0.76; p < 0.001). The use of the resection ratio of non-ecHFOs exhibited a higher AUC value of 0.82 (p < 0.001). The performance was further augmented by using a multiple regression model incorporating both the resection ratio of non-ecHFOs and complete removal of SOZ (AUC = 0.93, p =0.047).

**Figure 6:**
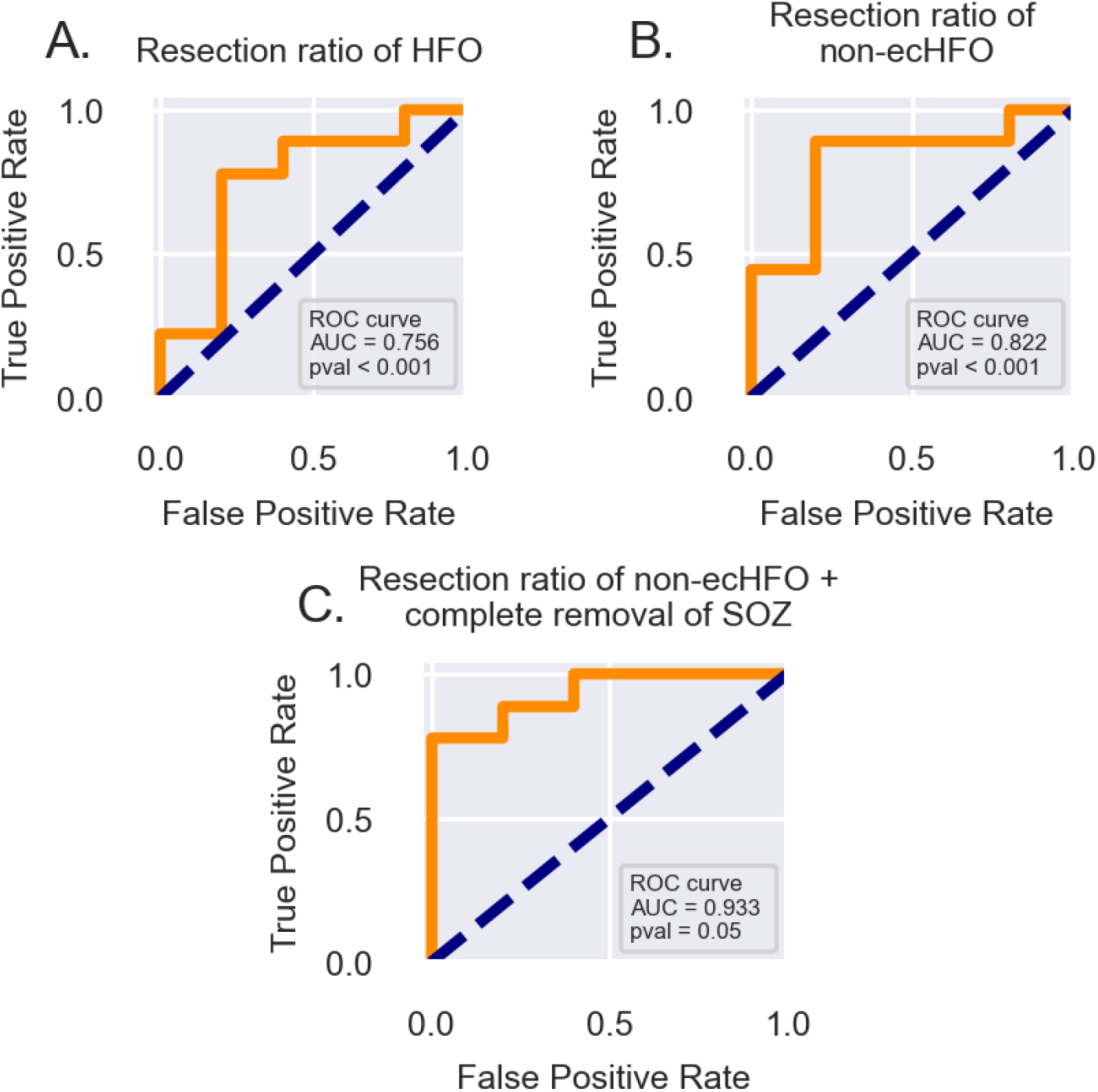
The accuracy of prediction models incorporating HFO resection ratio. We constructed postoperative seizure outcome prediction models using the HFO resection ratio derived from EEG data (n□=□14). Each receiver-operating characteristics (ROC) curve delineates the accuracy of seizure outcome classification of a given model, using the area under the ROC curve statistics. (A) Unclassified HFO resection ratio was used as a single classifier. (B) non-ecHFO resection ratio was used as a single classifier, which showed significant improvement in the prediction. (C) A multiple regression model incorporating the resection ratio of non-ecHFOs and complete removal of the SOZ (yes or no) was used, which demonstrated further improved predictive value of postoperative seizure outcomes. HFO = high-frequency oscillation; non-ecHFO = non-eloquent cortex HFOs; SOZ = Seizure onset zone.

## DISCUSSION

HFOs with similar frequency ranges emerge despite considerably different mechanisms.^35, 36^ A traditional hypothesis-driven approach to separate physiological HFOs from pathological HFOs poses challenges because numerous yet-to-be-identified features must be considered. Simple engineering features of HFOs, including amplitude, frequency, and duration, do not appear to successfully separate pathological from physiological HFOs.^37, 38^ Visual classification of HFOs with or without spikes along with artifact removal (such as ringing) is commonly performed because HFOs with spike-wave discharges are considered representative of pathological HFOs.^39^ However, this task is time-consuming and exhibits poor inter-rater reliability among human experts.^40, 41^ Fast ripples (250–500 Hz) might more specifically localize epileptogenic zones than ripples do (80–250 Hz), but their detection rate is much lower than ripples.^13^ Correcting the HFO detection rate with region-specific normative values seems a reasonable approach,^32, 42-44^ but this does not determine each HFO event as either pathological or physiological.

In the present study, we leveraged the robust clinical definition of the EC with cortical stimulation and gamma-related language mapping to identify cortical areas generating physiological HFOs (represented by ecHFOs). Our approach using a DL-based algorithm is distinct from other approaches. Based on our hypothesis that physiological HFOs look morphologically different from pathological HFOs, leveraging DL’s ability to analyze imaging input (transformed from EEG time-series data) seemed logical. By training a DL model with labels based on functional cortical mapping results. We then investigated the salient features of the physiological HFOs through the DL algorithm. The novel findings include that we found a bell-shaped template in the time-frequency plot as the discriminating feature of ecHFOs. More specifically, ecHFOs would have lower signal amplitude at the center of the HFO onset across the frequencies (including both ripple and fast ripple band) and lower signal amplitude in the low frequency band throughout the time window than non-ecHFOs (representing pathological HFOs). The ecHFOs generally had a slower peak frequency, a smaller amplitude, and tended to have a longer duration than non-ecHFOs. However, these time-domain characteristics showed significant overlaps between ecHFOs and non-ecHFOs, and none of such simple engineering features could clearly separate them, consistent with the previous studies.^37, 38^ Once the DL model was trained, we verified specific morphological features of ecHFOs via perturbation analysis. Insertion of spike templates significantly decreased ecHFO probability, which confirmed the traditional knowledge that HFOs with spikes are most likely pathological HFOs.

Our findings have significant clinical implications. Using the resection ratio (removed HFOs/detected HFOs) of non-ecHFOs (likely representing pathological HFOs), we demonstrated that the prediction of postoperative seizure outcomes significantly improved compared to using the uncorrected HFO resection ratio (AUC of 0.82, increased from 0.76). Combined with the current clinical standard of complete resection of SOZ, the AUC further improved to 0.93. The above findings suggest that the classification of HFOs (physiological vs. pathological) can potentially improve the clinical utility of HFOs in guiding resection, and this can still be combined with our current clinical standard of capturing habitual seizures to decide the resection margin.

A recent study demonstrated that a cortical location could produce different types of HFOs, such as both physiological and pathological HFOs.^19^ We probed this point by investigating cortical sites with both EC characteristics (behavioral responses with stimulation mapping or/and gamma activity with language tasks) and pathological properties (spontaneous spikes or/and AD/SZ). The ecHFO ratio was between cortical sites with only behavioral responses and that with spikes or/and SZ/AD only. The confidence scores of HFO type prediction are diffusely distributed from ecHFOs to non-ecHFOs, without clear bimodal distribution. If such “both” channels produce morphologically distinct physiological and pathological HFOs, one would expect to see a clear bimodal distribution. Our findings suggest that in cortical sites with both pathological and physiological properties, the morphologies of HFOs are heterogeneous. This may be a limitation of using macroelectrodes since multiple local generators might influence each HFO morphology.

There are several limitations in our study. We have analyzed only 18 patients to define physiological HFOs. Also, postoperative outcomes were correlated in only 14 patients. Although we analyzed an extended EEG dataset (90 minutes from each subject) to maximize the number of HFOs for analysis, we will need to analyze more patients to have a definitive conclusion. Although we assumed that HFOs from the healthy brain regions look morphologically similar, such characteristics may be different as a function of age and location of the brain regions. In addition, we analyzed only sleep EEG to maximize the number of HFOs for analysis. States of consciousness (including awake and different sleep stages) and vigilance levels may affect HFO morphology.^45^ With more number of subjects, we will be able to characterize the difference in ecHFOs generated in functionally distinct cortical areas, such as somatosensory, language, and visual cortex. Lastly, with an increasing number of diagnostic stereotactic EEG (SEEG) studies, it is of interest to investigate how HFO morphologies via SEEG would look different compared to strip/grid-sampled HFOs.

In summary, we proposed to use a weakly-supervised DL algorithm to characterize physiological HFOs using robust clinical outcomes (functional cortical mapping results) as training labels. Although future work to include a larger number of subjects will be needed, this method provided salient morphological features of physiological HFOs. Our results also suggested that this DL-based HFO classification, once trained, might help separate pathological from physiological HFOs, and efficiently guide surgical resection using HFOs.

## Data Availability

All data produced in the present study are available upon reasonable request to the authors

## ACKNOWLEDGMENT

The authors have no conflict of interest to disclose. HN is supported by the Sudha Neelakantan & Venky Harinarayan Charitable Fund, the Elsie and Isaac Fogelman Endowment, and the UCLA Children’s Discovery and Innovation Institute (CDI) Junior Faculty Career Development Grant (#CDI-SEED-010121; #CDI-TTCF-07012021). SAH has received research support from the Epilepsy Therapy Project, the Milken Family Foundation, the Hughes Family Foundation, the Elsie and Isaac Fogelman Endowment, Eisai, Lundbeck, Insys, Zogenix, GW Pharmaceuticals, UCB, and has received honoraria for service on the scientific advisory boards of Questcor, Mallinckrodt, Insys, UCB, and Upsher-Smith, for service as a consultant to Eisai, UCB, GW Pharmaceuticals, Insys, and Mallinckrodt, and for service on the speakers’ bureaus of Mallinckrodt and Greenwich Bioscience. RS serves on scientific advisory boards and speakers bureaus and has received honoraria and funding for travel from Eisai, Greenwich Biosciences, UCB Pharma, Sunovion, Supernus, Lundbeck Pharma, Liva Nova, and West Therapeutics (advisory only); receives royalties from the publication of Pellock’s Pediatric Neurology (Demos Publishing, 2016) and Epilepsy: Mechanisms, Models, and Translational Perspectives (CRC Press, 2011). RJS is supported by the National Institute of Neurological Disorders and Stroke (NINDS) R01NS106957. JEJ is supported by NINDS U54NS100064 and R01NS033310. The research described was also supported by NIH/National Center for Advancing Translational Science (NCATS) UCLA CTSI Grant Number UL1TR001881.

We are indebted to Jason T. Lerner, Lekha M. Rao, Rajsekar R. Rajaraman, Maria Garcia Roca, Richard Le, Patrick Wilson, and Jimmy C Nguyen for their assistance in the study and sample acquisition.

## AUTHOR CONTRIBUTIONS

We certify that all the authors listed made significant contributions to the work and share responsibility and accountability for the results.

- Conception and design of the study: YZ, WS, VR, HN
- Acquisition and analysis of data: YZ, HC, JN, TM, JHM, PW, MSS
- Drafting a significant portion of the manuscript or figures: YZ, SAH, JHM, AF, AE, RS, RJS, JE, WS, VR, HN

